# Denoising Strategies of Functional Connectivity MRI Data in Lesional and Non-Lesional Brain Diseases

**DOI:** 10.1101/2025.01.22.25320407

**Authors:** Stephan Wunderlich, Cagatay Alici, Saeed Motevalli, Veit Stoecklein, Louisa von Baumgarten, Florian Schöberl, Marion Subklewe, Enrico Schulz, Sophia Stoecklein

**Affiliations:** Department of Radiology, University Hospital Ludwig Maximilian University of Munich, Munich, Germany; School of Computation, Information, and Technology, Technical University of Munich, Munich, Germany; Department of Neurosurgery, University Hospital Ludwig Maximilian University of Munich, Munich, Germany; Department of Neurology, University Hospital Ludwig Maximilian University of Munich, Munich, Germany; Department of Medicine III, University Hospital Ludwig Maximilian University of Munich, Munich, Germany; German Cancer Consortium (DKTK), partner site Munich, German Cancer Research Center (DKFZ), Heidelberg, Germany; Department of Medicine III, University Hospital, Ludwig Maximilian University of Munich, Munich, Germany; Laboratory for Translational Cancer Immunology, Gene Center, Ludwig Maximilian University of Munich, Munich, Germany; Bavarian Cancer Research Center (BZKF), Erlangen, Germany

**Keywords:** Denoising Methods, FcMRI, Resting-State, Lesional, Non-lesional, Head Motion

## Abstract

**Purpose:** Functional connectivity magnetic resonance imaging (fcMRI) is a widely utilized tool for analyzing functional connectivity (FC) in both healthy and diseased brains. However, patients with brain disorders are particularly susceptible to head movement during scanning, which can introduce substantial noise and compromise data quality. Therefore, identifying optimal denoising strategies is essential to ensure reliable and accurate downstream data analysis for both lesional and non-lesional brain conditions.

**Approach:** In this study, we analyzed data from four cohorts: healthy subjects, patients with brain lesions (glioma, meningioma) and patients with a non-lesional encephalopathic condition. Our goal was to evaluate various denoising strategies using standard quality control (QC) metrics to identify the most effective approach for minimizing noise while preserving the integrity of the blood oxygen level-dependent (BOLD) signal, tailored to each disease type.

**Results:** The effectiveness of denoising strategies varied based on the data quality and whether the data was derived from lesional or non-lesional diseases. At comparable levels of head motion, combinations involving independent component analysis-based automatic removal of motion artifacts (ICA-AROMA) denoising strategies were most effective for data from an encephalopathic non-lesional condition, while combinations including anatomical component correction (CC) provided the best results for data from lesional conditions.

**Conclusion:** Here, we present the first comparison of denoising pipelines for patients with lesional and non-lesional brain diseases. A key finding was that, at comparable levels of head motion, the optimal denoising strategy varies depending on the nature of the brain disease.

## 1 Introduction

Resting-state functional connectivity magnetic resonance imaging (fcMRI) has become a crucial tool for investigating the brain’s connectional architecture in both healthy individuals and in patients with various brain pathologies (Ellis et al., 2023; Larrivee, 2024; St-Onge et al., 2023; Tai et al., 2023; Wei et al., 2024; Zhang et al., 2021). A critical aspect of fcMRI analysis is managing noise, as it can substantially distort measures of functional connectivity (FC) and complicate their interpretation (Van Dijk et al., 2012; Yan & Birn, 2023). This noise can arise from various sources, including physiological processes (e.g. cardiac pulsations, respiration), scanner-related artifacts (e.g. thermal noise, gradient imperfections), and subject-related factors like head motion (Liu, 2016). Among these, head motion is particularly problematic as it introduces spatial misalignment, spin history effects, and interactions with physiological signals, leading to spurious changes in the blood oxygen level-dependent (BOLD) signal (Maknojia et al. 2019). Studies have demonstrated that patients with brain pathologies, including brain tumors, are particularly prone to increased head motion during fcMRI scans (Seto et al., 2001; Silva et al., 2018). Also patients with conditions such as stroke, Alzheimer’s disease, bipolar disorder, and schizophrenia exhibit significantly greater head motion compared to age-matched healthy controls (Haller et al., 2014; Mowinckel et al., 2012; Wylie et al., 2014). Typically, in healthy cohorts, head motion ranges between 0.5 mm and 2 mm (Power et al., 2012; Van Dijk et al., 2012). However, in patients with brain diseases, head motion can often exceed these levels, reaching significantly higher values (Haller et al., 2014; Wu & Hallett, 2005). Depending on the study population, a significant proportion of data need to be excluded due to excessive head motion, e.g. approximately 25% in an imaging study involving newborn infants in spontaneous sleep (S. Stoecklein et al., 2020). High exclusion rates underscore the significant challenges of acquiring usable fcMRI data in clinical populations. The pervasive impact of head motion on data quality and the high prevalence of head motion artifacts in patient cohorts emphasize the critical need for robust denoising and head motion correction strategies to ensure accurate and reliable analysis of fcMRI data. Advanced denoising techniques can significantly enhance the usability of suboptimal data and improve overall outcomes (Parkes et al., 2018; Weiler et al., 2022). However, when data quality is severely compromised, it is essential to critically evaluate whether the dataset is appropriate for further analysis. Relying on excessively degraded data carries the risk of drawing inaccurate or misleading conclusions. Nonetheless, the clinical implementation of fcMRI must strive to ensure usability even in the presence of head motion artifacts, particularly in challenging patient populations, emphasizing the need for adaptive denoising pipelines (Lu et al., 2019).

Various denoising pipelines have been proposed in the literature for correcting noise, including independent component analysis-based automatic removal of head motion artifacts (ICA-AROMA) (Pruim et al., 2015), anatomical component correction (CC) (Behzadi et al., 2007), scrubbing (Power et al., 2012), head motion parameter (HMP) regression (Friston et al., 1996), Spike Regression (SpikeReg) (Cox, 1996), as well as regression approaches targeting global signal (GS) (Macey et al., 2004), cerebrospinal fluid (CSF), white matter (WM), and combinations of these methods. These denoising strategies have been widely used in the analysis of numerous fcMRI research questions and systematically evaluated in patients who were healthy or suffering from obsessive-compulsive disorder and schizophrenia (Ciric et al., 2017; Graff et al., 2022; Parkes et al., 2018) or in status post traumatic brain injury (Weiler et al., 2022). As outlined by Ciric and colleagues (Ciric et al., 2017) it is important to systematically evaluate denoising methods across diverse populations, as residual head motion artifacts may differ significantly between groups, potentially impacting the interpretability of FC results in clinical cohorts. Variability in pipeline effectiveness across groups underscores the importance of tailored approaches to address specific challenges, such as high head motion or anatomical distortions. Here, we aimed to systematically evaluate these methods in challenging patient populations to identify the optimal denoising strategies tailored to disease type and data quality. Our analysis included individuals exhibiting significant head motion during scanning as well as those with severe lesional brain conditions.

## 2 Methods

To determine the optimal denoising pipeline tailored to specific diseases, we evaluated commonly used denoising strategies in healthy subjects (n = 1000), in patients with lesional brain diseases (glioma (n = 34) and meningioma (n = 29)) and those with a non-lesional brain disease (encephalopathic condition (n = 14)). By assessing the performance of 16 widely applied denoising strategies (Table 3) (Parkes et al., 2018; Raval et al., 2020), we aimed to identify the most effective approach for each pathology type. This evaluation is based on three quality criteria to ensure a robust and comprehensive analysis of the pipelines’ suitability for both lesional and non-lesional brain conditions. The quality criteria are:

1. Quality control-functional connectivity (QC-FC) correlations (Ciric et al., 2017; Power et al., 2015)
2. QC-FC distance dependence (Ciric et al., 2017; Mahadevan et al., 2021; Power et al., 2015)
3. Loss in temporal degrees of freedom (tDOF-loss) (Ciric et al., 2017; Power et al., 2015)

### 2.1 Subjects

#### 2.1.1 Data from Healthy Subjects

We included 1000 patients (mean age 21.21 ± 2.69 years) (Table 1) from the Genomic Superstruct Project (GSP) (Holmes et al., 2015) fcMRI dataset as a healthy benchmark to evaluate the performance of the various denoising pipelines.

**Table 1:** Patient Characteristics. *Table 1 shows the number of patients and their characteristics, including their raw mFD. Genomics Superstruct Project (GSP), Mean Framewise Displacement (mFD), Immune Effector Cell-Associated Neurotoxicity Syndrome (ICANS), Standard Deviation (SD)*.

| **Cohorts** | **No. of Patients**<br>**[N]** | **Age**<br>**([years]**<br>**mean ± SD)** | **Sex**<br>**([N]**<br>**F/M)** | **mFD**<br>**([mm]**<br>**mean ± SD)** |
| --- | --- | --- | --- | --- |
| **GSP** |  |  |  |  |
| No-Exclusion | 1000 | 21.21 ± 2.69 | 550/450 | 0.182 ± 0.077 |
| Medium-level Motion | 0 | - | - | - |
| High-level Motion | 0 | - | - | - |
| **ICANS** |  |  |  |  |
| No-Exclusion | 14 | 62.60 ± 15.38 | 7/7 | 0.534 ± 0.137 |
| Medium-level Motion | 6 | 60.83 ± 11.02 | 2/4 | 0.421 ± 0.063 |
| High-level Motion | 8 | 65.75 ± 08.55 | 4/4 | 0.619 ± 0.110 |
| **Glioma** |  |  |  |  |
| No-Exclusion | 34 | 48.70 ± 16.90 | 14/20 | 0.340 ± 0.087 |
| Medium-level Motion | 24 | 48.00 ± 16.65 | 14/10 | 0.390 ± 0.042 |
| High-level Motion | 0 | - | - | - |
| **Meningioma** |  |  |  |  |
| No-Exclusion | 29 | 58.20 ± 12.30 | 22/7 | 0.336 ± 0.067 |
| Medium-level Motion | 17 | 61.23 ± 07.98 | 10/7 | 0.383 ± 0.043 |
| High-level Motion | 0 | - | - | - |

#### 2.1.2 Data from Lesional Conditions

For intra-axial data from lesional conditions, we included patients with glioma, and for extra-axial data from lesional conditions, we used patients with meningioma. These data were reanalyzed from a previous study (V. M. Stoecklein et al., 2020), which included 34 patients with glioma (mean age = 48.7 ± 16.9 years) (Table 1). Data from 29 patients with meningioma (mean age = 58.2 ± 12.3 years) (V. M. Stoecklein et al., 2023), were also reanalyzed for this research (Table 1). Histopathological diagnoses were obtained according to the 2016 World Health Organization classification criteria (Louis et al., 2016). The studies were reviewed and approved by the local institutional review board of Ludwig Maximilian University of Munich. All study procedures were in accordance with the Declaration of Helsinki. Written informed consent was obtained from all patients (Approval Number 676-15).

#### 2.1.3 Data from a Non-Lesional Condition

This cohort included 14 patients (mean age = 62.6 ± 15.4 years), with lymphoma or melanoma who developed symptoms of immune effector cell-associated neurotoxicity syndrome (ICANS) during chimeric antigen receptor (CAR)-T-cell therapy (Table 1). Data were partially included in a previous publication (S. Stoecklein et al., 2023). All data from patients undergoing CAR-T-cell therapy were collected during ICANS, in some cases with significant head movement (Figure 1). Despite their neurotoxic symptoms, their routine clinical magnetic resonance imaging (MRI) exams, including contrast-enhanced sequences, were unremarkable. The study was reviewed and approved by the local institutional review board of Ludwig Maximilian University Munich. All study procedures were in accordance with the Declaration of Helsinki. Written informed consent was obtained from all patients (Approval Number: 646-20).

**Figure 1:**
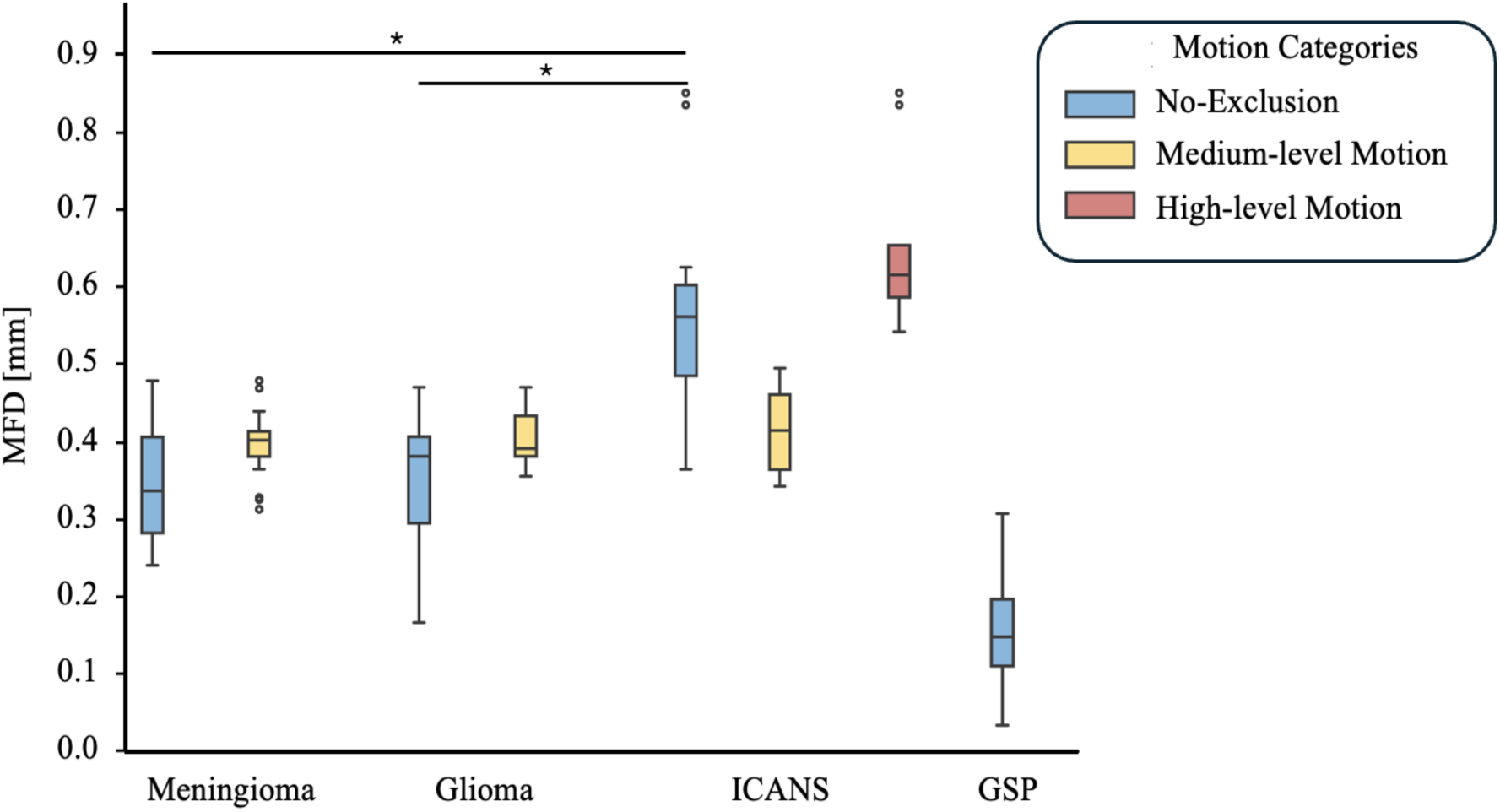
Raw Mean Framewise Displacement. *Figure 1 depicts the mFD values across different patient cohorts, categorized by motion categories: No-exclusion (all mFD values), medium-level motion (mFD between 0.3 and 0.5 mm) and high-level motion (mFD > 0.5 mm). The GSP dataset exhibits the lowest mFD values, followed by the no-exclusion category for Meningioma and Glioma. The highest raw mFD values are observed in the high-motion category among ICANS patients. Significant differences (p < 0.05) between patient cohorts (Meningioma, Glioma, ICANS) are indicated with an asterisk (*). Notably, no significant differences were observed among the three patient cohorts in the medium-level motion category. Genomics Superstruct Project (GSP), Mean Framewise Displacement (mFD), Immune Effector Cell-Associated Neurotoxicity Syndrome (ICANS)*.

### 2.2 Scanning Procedure

FcMRI data for the three patient cohorts followed the same scanning protocol. The data were collected on a 3T Skyra scanner (Siemens Medical Solutions) using a 20-channel head and neck coil. Functional images were acquired using an echo planar imaging pulse sequence (repetition time = 3000 ms, echo time = 30 ms, flip angle = 90°, 3 × 3 × 3 mm3 voxels, 120 time points per run). Each participant underwent two resting-state runs of six minutes each, resulting in a total of 240 time points. Structural MRI scans were acquired using a sagittal 3D T1-weighted sequence (radiofrequency pulses and magnetization prepared rapid acquisition gradient echo (MPRAGE) sequence, 1 × 1 × 1 mm3 voxels). The GSP scanning protocol is largely consistent with the scanning protocol above, except for the use of 3T Tim Trio scanners with a flip angle of 80° and an MPRAGE sequence resolution of 1.2 × 1.2 × 1.2 mm³ voxels.

### 2.3 Data Preprocessing

Structural MRI data were processed using the FreeSurfer version 6.0 software package (http://surfer.nmr.mgh.harvard.edu). Preprocessing of the fcMRI datasets was performed using FMRIPREP version 20.2.2, based on Nipype (Esteban et al., 2019, 2020; Gorgolewski et al., 2011). The first five functional images were deleted. The remaining frames were standardized, detrended and smoothed with a 6.0 mm full width at half maximum Gaussian kernel. The BOLD signal was band-pass filtered (0.01 Hz to 0.08 Hz). Finally, Power atlas containing 264 ROIs was used for brain parcellation (Power et al., 2011).

### 2.4 Data Grouping

We stratified patient groups (Table 1), based on the mean framewise displacement (mFD) of the raw functional data. We defined three motion categories (Table 2, Figure 1). The first category encompassed all data, while the second comprised data with an mFD falling between 0.3 mm and 0.5 mm (Aquino et al., 2019; Power et al., 2014; Smith et al., 2022). The third motion category included data with a mFD exceeding 0.5 mm, observed exclusively in patients from the ICANS cohort.

**Table 2:** Motion Category and their Criteria for Exclusion. *Table 2 outlines the criteria for defining the different motion categories based on raw mFD values. Mean Framewise Displacement (mFD)*.

| **Motion Category** | **Exclusion Criteria** |
| --- | --- |
| No-Exclusion | - |
| Medium-level Motion | Excluded subjects if:<br>(i) $mFD \geq 0.5$ mm; or<br>(ii) $mFD < 0.3$ mm. |
| High-level Motion | Excluded subjects if:<br>(i) $mFD < 0.5$ mm. |

### 2.5 Denoising Pipelines

The selected denoising strategies (Table 3) are grounded in established approaches from the literature and encompassed commonly used denoising methods to effectively reduce noise. The combinations were chosen based on their frequent application in previous studies and their ability to leverage the strengths of individual methods. This design enabled a comprehensive comparison of their performance and interactions, providing valuable insights into their effectiveness (Golestani & Chen, 2024; Parkes et al., 2018; Wei et al., 2024). All denoising procedures and their combinations were systematically applied to the four cohorts.

**Table 3:** Overview of the Different Denoising Pipelines. *Table 3 displays the different denoising strategies that were evaluated. Head Motion Parameters (HMP), Independent Component Analysis-based Automatic Removal of Motion Artifacts (ICA), Anatomical Component Correction (CC), Spike Regression (SpikeReg), Scrubbing (Scr)*.

| Pipeline | Denoising Strategies |
| --- | --- |
| 1 | 6HMP |
| 2 | 24HMP |
| 3 | SpikeReg |
| 4 | Scr |
| 5 | ICA |
| 6 | CC |
| 7 | ICA + 24HMP |
| 8 | ICA + SpikeReg |
| 9 | ICA + Scr |
| 10 | CC + 24HMP |
| 11 | CC + SpikeReg |
| 12 | CC + Scr |
| 13 | ICA + SpikeReg + 24HMP |
| 14 | ICA + Scr + 24HMP |
| 15 | CC + SpikeReg + 24HMP |
| 16 | CC + Scr + 24HMP |

Regression of WM, CSF and GS as calculated in FMRIPREP version 20.2.2 were consistently included throughout all denoising strategies. Additionally, the choice not to combine ICA-AROMA and CC within the same pipeline was intentional and based on methodological considerations. ICA-AROMA uses independent component analysis to identify and remove noise components, particularly head motion-related artifacts, based on their spatial and temporal characteristics. In contrast, CC utilizes principal component analysis to extract noise components from physiological sources, such as WM and CSF. While these methods are effective independently, combining them could introduce redundancy, potentially leading to excessive signal removal or diminished interpretability of results (Artoni et al., 2018).

#### 2.5.1 Regression of Head Movement Parameters (HMP)

We applied a 6 HMP model to correct for six translational and rotational head movement dimensions. Additionally, an extended model with 24 HMP, including their derivatives, was used (Satterthwaite et al., 2013).

#### 2.5.2 Anatomical Component Correction (CC)

The CC method models noise in BOLD signals using anatomical principal component analysis (Behzadi et al., 2007; Muschelli et al., 2014). CC regresses principal components from WM and CSF to account for non-neuronal origins. We extracted always the leading 10 confound regressors.

#### 2.5.3 Independent Component Analysis-based Automatic Removal of Motion Artifacts (ICA-AROMA)

ICA-AROMA, a method developed by (Pruim et al., 2015) based on independent component analysis, classifies spatially independent components (ICs) as either noise or neural signals using predefined criteria. Users have the option to specify a fixed number of ICs for removal or utilize an automatic estimator. According to (Pruim et al., 2015), the automatic method is recommended when the number of ICs is below 100, as was the case in our project, where we opted for the automatic version.

#### 2.5.4 Scrubbing and Spike Regression (SpikeReg)

In scrubbing, volumes were identified as contaminated and removed based on framewise displacement (FD) (Muschelli et al., 2014; Power et al., 2012). Additionally, we applied SpikeReg (Satterthwaite et al., 2013), where a separate regressor was generated for each volume with excessive head motion. In both methods, a threshold of FD > 0.3 mm was used to identify volumes affected by head motion.

### 2.6 Quality Control (QC) Measures

To evaluate the denoising strategies, the following quality control (QC) measurements were used:

MFD: MFD measures the mean movement between consecutive volumes (frames) of fcMRI data. The mFD was calculated by summing the absolute values of the derivatives of the six realignment parameters obtained from head motion correction algorithms, representing translation and rotation movements (Power et al., 2012). Higher mFD values indicate greater head motion during the scan, which can introduce noise and artifacts into the data and potentially confound FC analysis. We calculated the mFD for the raw data.

TDOF-loss: The quantity of time points in a BOLD dataset determines the degrees of freedom for statistical inference, with fewer points potentially leading to increased FC (Yan et al., 2013). TDOF-loss describes a reduction in the effective number of independent observations in the temporal dimension of fcMRI data due to preprocessing. Maintaining an adequate number of temporal degrees of freedom is critical for precise estimation of statistical significance. Decreased temporal degrees of freedom can result in inflated false positive rates or reduced sensitivity in detecting genuine neural activity. Therefore, it is crucial to strike a balance between head motion correction and preserving sufficient tDOF. We calculated tDOF-loss after applying each denoising strategy.

QC-FC Correlation: This measurement explores the relationship between FC and mFD, as non-neuronal fluctuations can inflate FC measurements by introducing erroneous shared variability across time series (Ciric et al., 2017). We computed Pearson’s correlation coefficient between FC values of each pair of our defined regions of interest (ROIs) and the corresponding mFD across all patients. Next, we analyzed the ratio of ROIs where the correlation between QC and FC was statistically significant (p < 0.05) after the application of each denoising pipeline. A higher QC-FC score indicates a denoising pipeline’s inefficacy in noise mitigation within FC.

QC-FC Distance Dependence: The QC-FC distance-dependence explores how mFD varies concerning the spatial separation between brain regions, aiming to discern whether mFD’s effectiveness in reducing FC artifacts is contingent on the distance between analyzed regions. While some QC measures may excel in mitigating FC artifacts for closely situated regions, their efficacy might diminish as spatial distance increases (Power et al., 2012, 2015; Van Dijk et al., 2012). Understanding this dependence is pivotal for assessing FC estimate reliability and refining QC procedures to adapt to spatial variability in artifact correction. We computed the distance between our defined ROI pairs by calculating their Euclidean coordinates and then evaluated the correlation between this distance measure and the QC-FC correlation for each edge using Spearman’s rank correlation coefficient (*ρ*), ideal for capturing nonlinear associations between variables.

### 2.7 Composite Score

The composite score combines the three QC metrics - QC-FC, QC-FC distance dependence, and tDOF-loss - using weighted (0.4, 0.4, 0.2) normalization to provide an integrated evaluation of the different denoising pipeline performance. By assigning tailored weights to each metric, the composite score balances the impact of head motion artifacts, spatial biases, and degrees of freedom, ensuring a comprehensive assessment of data quality. We prioritized QC-FC metrics which are critical for ensuring the validity of FC estimates (Power et al., 2012).

### 2.8 Statistical Analysis

To test for significant differences in the raw mFD values between the different patient cohorts for the no-exclusion and medium-level motion categories, we used first the Shapiro-Wilk-Test to test for normal distribution. Given non-normal distributions, we applied the Kruskal-Wallis test to evaluate differences among groups. After the Kruskal-Wallis test (significant if p < 0.05), we conducted post-hoc pairwise comparisons using the Mann-Whitney U test (significant if p < 0.05). To mitigate the risk of false positives due to multiple comparisons, we applied the Bonferroni correction, adjusting the significance threshold based on the number of pairwise tests.

## 3 Results

### 3.1 Raw Mean Framewise Displacement (mFD)

Figure 1 presents the baseline mFD values across the different disease types and motion categories. The GSP dataset exhibited the lowest mean mFD 0.182 ± 0.077 (median = 0.164), indicating minimal head motion during scanning. This was followed by participants with lesional pathologies. The highest mFD values were observed in patients in ICANS, reflecting the greatest degree of head motion among all groups. In the no-exclusion category, mean mFD was 0.336 ± 0.067 (median = 0.320) for patients with meningioma (N = 29), 0.340 ± 0.087 (median = 0.369) for patients with glioma (N = 34), and 0.534 ± 0.137 (median = 0.537) for patients in ICANS (N = 14). For the medium-level motion category, patients with meningioma had a mean mFD of 0.383 ± 0.043 (median = 0.389) (N = 17), patients with glioma had a mean mFD of 0.390 ± 0.042 (median = 0.378) (N = 24), and patients in ICANS had a mean mFD of 0.421 ± 0.063 (median = 0.414) (N = 6). Lastly, in the high-level motion category, patients in ICANS had a mean mFD of 0.619 ± 0.110 (median = 0.572) (N = 8). The distributions of all patient cohorts were non-normal. In the no-exclusion category patients in ICANS showed significantly higher mFD than patients with meningioma (p < 0.01), and patients with glioma (p < 0.01). No significant difference was observed between patients with meningioma and with glioma (p > 0.2). For the medium-level motion category, no significant differences in mFD between the three patient groups were detected (all p-values > 0.2).

### 3.2 Quality Control-Functional Connectivity Correlation (QC-FC)

When evaluating the correlation between head motion and FC, we found that the combination of CC + SpikeReg + 24HMP yielded the best performance for data from lesional conditions (Figure 2, 3) across all motion categories. Conversely, ICA + SpikeReg + 24HMP was most successful for data from the non-lesional condition (Figure 2, 3). Healthy patients (GSP) demonstrated the best overall results, with a slightly superior performance observed for the combination of CC + SpikeReg + 24HMP compared to ICA + SpikeReg + 24HMP (Figure 2).

**Figure 2:**
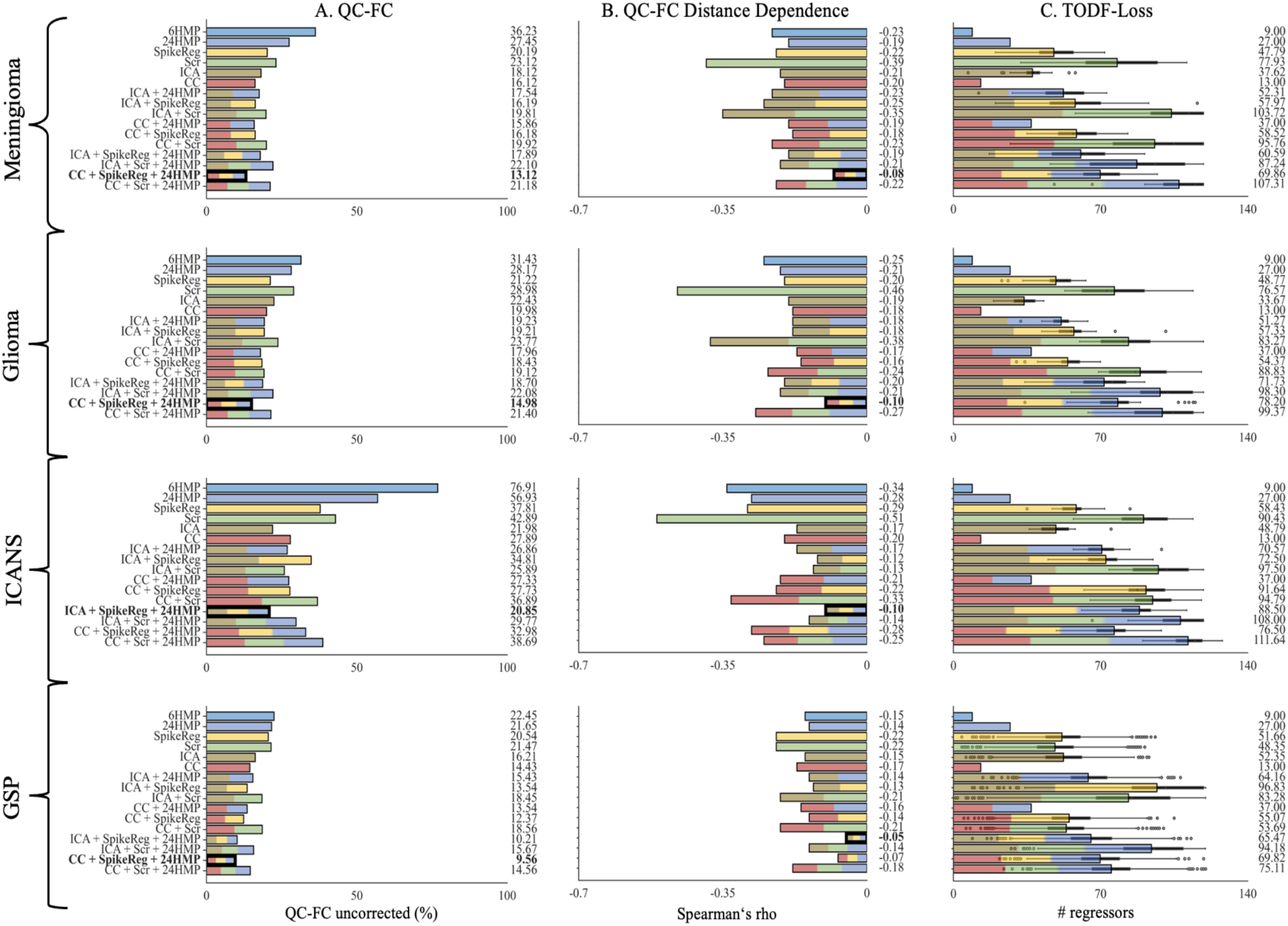
Quality Control – Functional Connectivity (QC-FC), QC-FC Distance Dependence and Loss of Temporal Degree of Freedom Metrics in the No-Exclusion Category. *This figure illustrates the performance of the denoising strategies applied across all patients for A. QC-FC, B. QC-FC Distance Dependence and C. TODF-Loss. Each denoising method is represented by a distinct color, while pipelines that combine multiple denoising approaches are depicted as bars with evenly distributed segments in the corresponding colors. The best-performing pipelines are highlighted in bold (for QC-FC and QC-FC distance dependence only), with lower correlation values indicating superior performance. For QC-FC metrics, the **CC + SpikeReg + 24HMP** pipeline emerges as the top-performing strategy for data from **lesional** conditions, while the **ICA + SpikeReg + 24HMP** pipeline proves to be the best for data from the **non-lesional** condition. In terms of tDOF-loss, it is logically lowest in simpler pipelines such as HMP and CC. However, among the identified best-performing pipelines, a notable difference emerges: In data from lesional conditions, the ICA + SpikeReg + 24HMP pipeline demonstrates lower TDOF-loss compared to CC + SpikeReg + 24HMP, whereas in data from the non-lesional condition, the opposite holds true. Quality Control (QC), Quality Control - Functional Connectivity (QC-FC), Genomics Superstruct Project (GSP), Global Signal Regression (GSR), Head Motion Parameter (HMP), Spike Regression (SpikeReg), Scrubbing (Scr), Independent Component Analysis-based Automatic Removal of Motion Artifacts (ICA), Anatomical Component Correction (CC)*.

**Figure 3:**
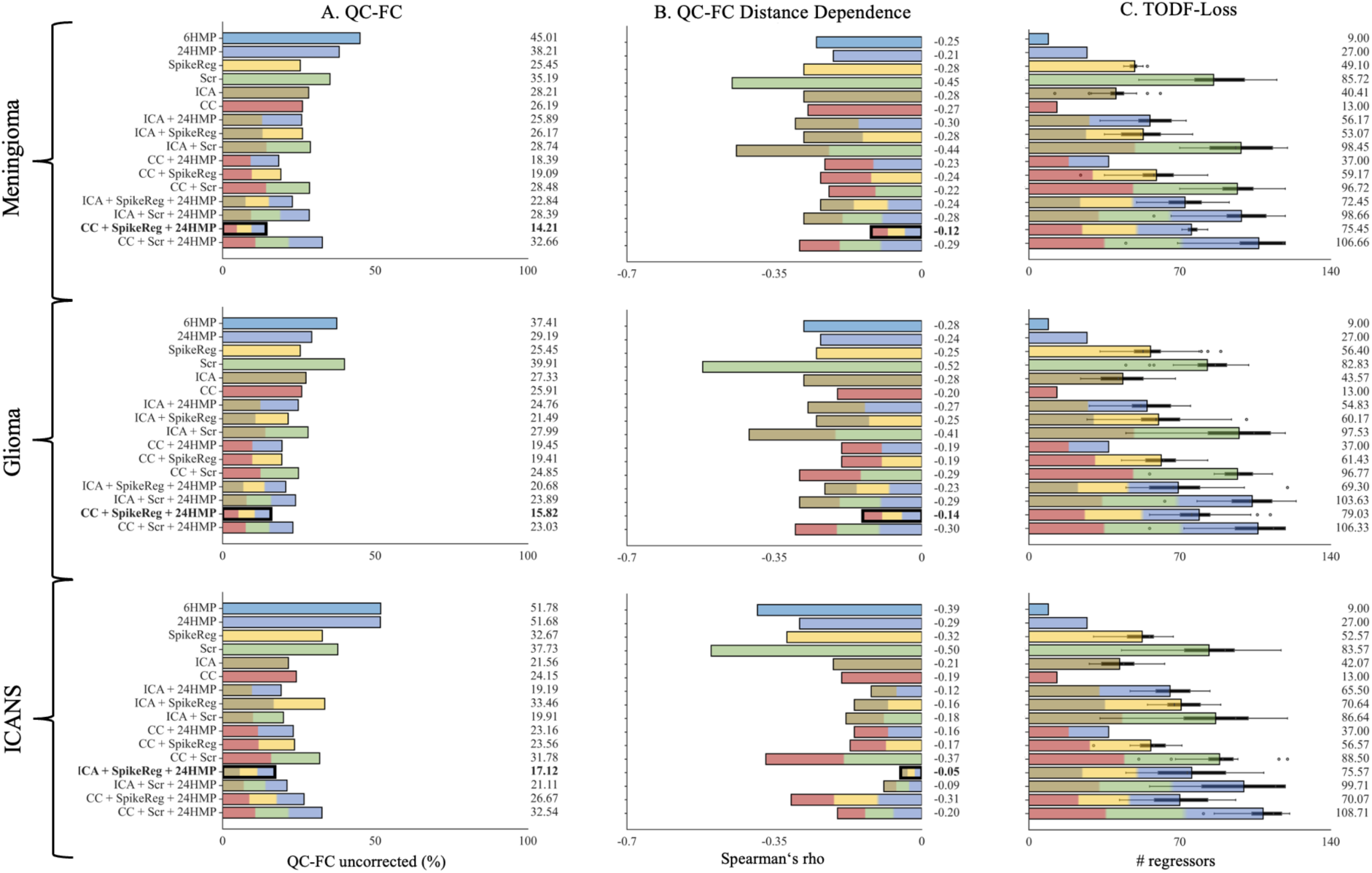
Quality Control – Functional Connectivity (QC-FC), QC-FC Distance Dependence and Loss of Temporal Degree of Freedom Metrics in the Medium-Level Motion Category. *The figure illustrates the performance of the denoising strategies in the medium-level motion category (mFD from 0.3 to 0.5mm) for A. QC-FC, B. QC-FC Distance Dependence and C. TODF-Loss. Each denoising method is represented by a distinct color, while pipelines that combine multiple denoising approaches are depicted as bars with evenly distributed segments in the corresponding colors. The best-performing denoising pipelines are highlighted in bold (for QC-FC and QC-FC distance dependence only), with strategies yielding the lowest correlations being preferred. For QC-FC metrics, the **CC** + **SpikeReg** + **24HMP** pipeline emerges as the top-performing strategy for data from **lesional** conditions, while the **ICA** + **SpikeReg** + **24HMP** pipeline proves to be the best for data from the **non-lesional** condition. In terms of tDOF-loss, it is logically lowest in simpler pipelines such as HMP and CC. However, among the identified best-performing pipelines, a notable difference emerges: In data from lesional conditions, the ICA + SpikeReg + 24HMP pipeline exhibits lower TDOF-loss than CC + SpikeReg + 24HMP, whereas in data from the non-lesional condition, the reverse is true. Quality Control (QC), Quality Control - Functional Connectivity (QC-FC), Global Signal Regression (GSR), Head Motion Parameter (HMP), Spike Regression (SpikeReg), Scrubbing (Scr), Independent Component Analysis-based Automatic Removal of Motion Artifacts (ICA), Anatomical Component Correction (CC)*.

### 3.3 Quality Control-Functional Connectivity Correlation (QC-FC) Distance Dependence

When considering brain region distance, the correlation between head motion and FC exhibited similar patterns to the analysis without distance dependence. Concretely, CC + SpikeReg + 24HMP demonstrated the best performance in data from lesional conditions. For data from the non-lesional condition, ICA + SpikeReg + 24HMP proved to be the most effective. Healthy patients (GSP) demonstrated the best overall results, with a slightly superior performance observed for the combination of ICA + SpikeReg + 24HMP compared to CC + SpikeReg + 24HMP (Figure 2, 3).

### 3.4 Loss of Temporal Degree of Freedom (tDOF-loss)

Surprisingly, in tDOF-loss we observed the contrary performance as in the other QC methods for data from lesional and non-lesional conditions. Combinations involving ICA-AROMA again ICA-AROMA (ICA) + SpikeReg + 24HMP, performed better in preserving the tDOF in data from lesional conditions compared to CC + SpikeReg + 24HMP. For patients in ICANS, ICA + SpikeReg + 24HMP showed slightly worse tDOF preservation than CC + SpikeReg + 24HMP in both motion categories (Figure 2, 3). Logically, the lowest loss is seen in 6 HMP and CC alone.

### 3.5 High-Level Motion Data in Patients in ICANS

In this sub-cohort, we focused exclusively on patients with the non-lesional condition with high-level motion to evaluate the performance of denoising methods under very noisy conditions. Consistent with previous sections, the combination of ICA + SpikeReg + 24HMP emerged as the best-performing pipeline for data from the non-lesional condition in QC-FC and QC-FC distance-dependence metrics. Furthermore, also here tDOF-loss is lower for CC + SpikeReg + 24HMP compared to ICA + SpikeReg + 24HMP as can be seen in Figure 4.

**Figure 4:**
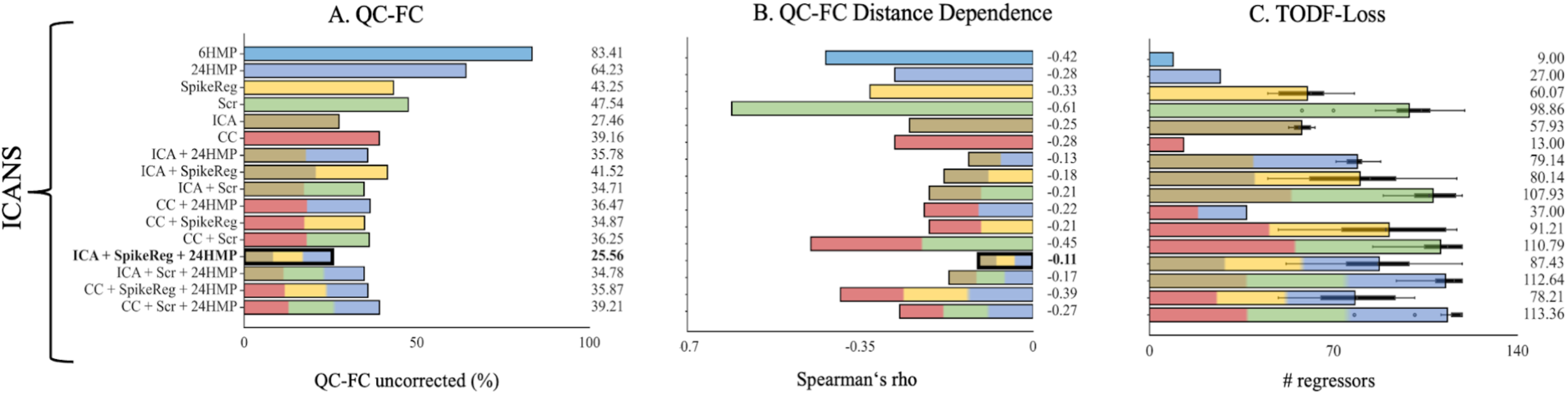
Quality Control – Functional Connectivity (QC-FC), QC-FC Distance Dependence and Loss of Temporal Degree of Freedom Metrics in patients with ICANS in the High-Level Motion Category. *The figure illustrates the performance of the denoising strategies within the high-motion category (mFD > 0.5 mm) for A. QC-FC, B. QC-FC Distance Dependence and C. TODF-Loss, focusing exclusively on patients from the ICANS cohort. Each denoising method is represented by a distinct color, while pipelines that combine multiple denoising approaches are depicted as bars with evenly distributed segments in the corresponding colors. The best-performing denoising pipelines are highlighted in bold (for QC-FC and QC-FC distance dependence only), with strategies yielding the lowest correlations being preferred. The **ICA + SpikeReg + 24HMP** pipeline proves to be the best for the QC-FC metrics. In terms of tDOF-loss, it is logically lowest in simpler pipelines such as HMP and CC. However, the **CC + SpikeReg + 24HMP** exhibited lower loss of tDOF compared to **ICA + SpikeReg + 24HMP**. Quality Control - Functional Connectivity (QC-FC), Global Signal Regression (GSR), Head Motion Parameter (HMP), Spike Regression (SpikeReg), Scrubbing (Scr), Independent Component Analysis-based Automatic Removal of Motion Artifacts (ICA), Anatomical Component Correction (CC)*.

### 3.6 Data from Lesional Conditions

Interestingly, the combination of CC + SpikeReg + 24HMP yielded similarly favorable results for both patients with meningioma and glioma. Furthermore, across QC metrics, patients with meningioma tend to achieve slightly better outcomes with the optimal denoising method (QC-FC: 13.12, QC-FC distance dependence: -0.08) compared to patients with glioma (QC-FC: 14.98, QC-FC distance dependence: -0.10).

### 3.7 Composite Score

To provide a comprehensive and overarching interpretation of the various QC metrics, we calculated a composite score for each patient cohort, motion category and denoising pipeline (Table 4). A lower composite score indicates better pipeline performance across the three QC metrics compared to other pipelines within the same patient cohort and motion category. For patients in ICANS, ICA and combinations, especially ICA + SpikeReg + 24HMP demonstrated the best performance (composite score for the medium-level motion category = 0.178). In data from lesional conditions, CC and its combinations, particularly CC + SpikeReg + 24HMP, achieved the best results (composite score for the medium-level motion category in patients with glioma = 0.246, in patients with meningioma = 0.234). In the GSP dataset best was ICA + SpikeReg + 24HMP (composite score = 0.239), but closely followed by CC + SpikeReg + 24HMP (composite score = 0.266). It is important to state that each composite value does not indicate the best denoising pipeline through all patient cohorts and motion categories, but only the best denoising pipeline within each pathology and motion category.

**Table 4:** Composite Scores of Denoising Pipelines. *Table 4 presents for every pathology and motion category the top two pipelines ranked by their composite scores, which are calculated using QC-FC, QC-FC distance dependence, and tDOF Loss metrics with respective weights of 0.4, 0.4, and 0.2. Genomics Superstruct Project (GSP), Head Motion Parameter (HMP), Spike Regression (SpikeReg), Scrubbing (Scr), Independent Component Analysis-based Automatic Removal of Motion Artifacts (ICA), Anatomical Component Correction (CC)*.

|  | Denoising Pipeline | Composite Score |
| --- | --- | --- |
| **ICANS** |  |  |
| 1. No-Exclusion | ICA | 0.210 |
| 1. No-Exclusion | ICA + SpikeReg + 24HMP | 0.210 |
| 1. Medium-level Motion | ICA + SpikeReg + 24HMP | 0.178 |
| 2. Medium-level Motion | CC | 0.242 |
| 1. High-level Motion | ICA + SpikeReg + 24HMP | 0.219 |
| 2. High-level Motion | ICA | 0.271 |
| **Glioma** |  |  |
| 1. No-Exclusion | CC + SpikeReg + 24HMP | 0.225 |
| 2. No-Exclusion | CC + 24HMP | 0.276 |
| 1. Medium-level Motion | CC + SpikeReg + 24HMP | 0.246 |
| 2. Medium-level Motion | CC + 24HMP | 0.262 |
| **Meningioma** |  |  |
| 1. No-Exclusion | CC + SpikeReg + 24HMP | 0.209 |
| 2. No-Exclusion | CC | 0.265 |
| 1. Medium-level Motion | CC + SpikeReg + 24HMP | 0.234 |
| 2. Medium-level Motion | CC + 24HMP | 0.313 |
| **GSP** |  |  |
| 1. No-Exclusion | ICA + SpikeReg + 24HMP | 0.239 |
| 2. No-Exclusion | CC + SpikeReg + 24HMP | 0.266 |

## 4 Discussion

In this study, we provide a comprehensive comparison of various fcMRI denoising strategies applied to data from healthy subjects, patients with lesional brain disorders (glioma and meningioma), and those with a non-lesional encephalopathic condition, ICANS. Each strategy was rigorously evaluated for its ability to minimize noise interference in fcMRI data using established QC parameters (Parkes et al., 2018). QC-FC analyses underscored the effectiveness of ICA + SpikeReg + 24HMP for data from the non-lesional condition and while CC + SpikeReg + 24HMP performed best for data from lesional conditions. Composite scores of QC parameters confirmed these pipelines as the most effective for their respective data types.

Across all motion categories in the non-lesional condition, ICA-AROMA-based combinations, particularly ICA + SpikeReg + 24HMP, excelled in reducing both QC-FC and QC-FC distance dependence. This aligns with the findings of (Parkes et al., 2018), who similarly demonstrated the efficacy of denoising strategies involving ICA-AROMA across their entire dataset of non-lesional conditions, including both healthy subjects and patients, regardless of head motion levels. However, these results contrast with (Weiler et al., 2022), who found that ICA-AROMA was ineffective to reduce noise in patients with status post traumatic brain injury. In data from lesional conditions, denoising strategies with ICA-AROMA did not perform well in reducing QC-FC and QC-FC distance dependence. Instead, the combination CC + SpikeReg + 24HMP performed better in data from lesional conditions. The effectiveness of ICA + SpikeReg + 24HMP in data from the non-lesional condition and CC + SpikeReg + 24HMP in data from lesional conditions became more evident when considering medium-level motion data, which likely represents the majority of real-world scanned data. Even in high-level motion data in patients in ICANS, combinations with ICA-AROMA still showed a significant reduction in QC-FC and QC-FC distance dependence. Since SpikeReg and 24HMP are common to both best-performing denoising pipelines (ICA + SpikeReg + 24HMP and CC + SpikeReg + 24HMP), their combined use appears to be beneficial for both disease types. This suggests that the differing effectiveness of ICA-AROMA and CC in data from lesional versus from non-lesional conditions may be attributed to underlying factors unique to each dataset.

A possible reason ICA-AROMA performs poorly at reducing QC-FC and QC-FC distance dependence in data from lesional conditions, even though it has a low tDOF loss, is that it struggles to distinguish real neural signals from noise in these cases. This challenge may originate from the intrinsic variability in frequency distributions observed in brain data impacted by lesions, particularly in regions affected by tumors and their immediate surroundings. Moreover, in patients with glioma, regions distal to the actual tumor site can also exhibit significant variability in the BOLD signal (Stoecklein et al. 2020; Osswald et al. 2016; Salvalaggio et al. 2024; Hadjiabadi et al. 2018) and therefore also in its frequency distributions. Specifically, there appears to be a pronounced shift in the BOLD signal towards lower frequencies in data from lesional conditions. This tendency towards lower frequency oscillations may reflect underlying changes in neural connectivity or activity associated with the lesion (Agarwal et al., 2023; Falcó-Roget et al., 2024; Urbin et al., 2014). The shift in the BOLD signal towards lower frequencies in data from lesional conditions may not only reflect changes in neural connectivity or activity but could also be influenced by perfusion and vascular effects associated with the lesion. Brain lesions can disrupt normal cerebral blood flow and vascular reactivity, leading to alterations in the hemodynamic response underlying the BOLD signal. For instance, gliomas exhibit areas of disrupted neurovascular coupling, where the relationship between neural activity and blood flow is impaired (Agarwal et al., 2023; Pak et al., 2017). Additionally, perifocal edema and tumor-related vascular remodeling can contribute to these effects by altering baseline perfusion and creating regions of hypoperfusion or hyperperfusion (Osswald et al., 2016; Rieger & Welter, 2015). This variability might pose challenges for ICA-AROMA’s ability to accurately distinguish between true neural activity and noise in data from lesional conditions. ICA-AROMA identifies motion-related ICs based on different criteria, where one is if the BOLD signal contains more than 35% high-frequency content based on a threshold calculated within the ICA-AROMA procedure (Pruim et al., 2015). As a result, this shift frequently gives rise to a greater number of ICs characterized by lower frequencies. This phenomenon can be described by the intrinsic properties of the BOLD signal: when its frequency diminishes, the resultant ICs derived from it tend to mirror these lower frequencies. This correlation arises from the nature of ICA, which dissects the signal into components that uphold the temporal traits of the original data. Despite this, these ICs may still contain noise, although they meet the 35% criteria. Consequently, ICA-AROMA might struggle to accurately distinguish between motion-affected ICs and non-motion-affected ICs in data from lesional conditions and classify fewer ICs as motion-affected. This is evident from the limited reduction in QC-FC and QC-FC distance dependence in data from lesional conditions, as well as the unexpectedly low tDOF loss observed with pipelines incorporating ICA-AROMA in these cases.

CC is based on a principal component analysis and focuses on removing noise components based on anatomical regions like WM and CSF (Behzadi et al., 2007). In data from lesional conditions, these regions might be more clearly defined in terms of noise characteristics, allowing CC to effectively remove noise specific to these areas defined in our case by the tumor. This hypothesis is further supported by the enhanced efficacy of CC + SpikeReg + 24HMP in patients with meningioma compared to those with glioma. This difference in response may be attributed to the distinct biological characteristics of these tumors. Meningiomas typically manifest as well-defined, localized lesions, whereas gliomas often present with diffuse, bilateral alterations in the BOLD signal (Stoecklein et al. 2020; Osswald et al. 2016; Salvalaggio et al. 2024; Hadjiabadi et al. 2018). As a result, denoising pipelines may find it inherently more manageable to address the specific and localized changes associated with meningiomas.

The integrated evaluation using a composite score reveals that the same top pipelines - CC + SpikeReg + 24HMP and ICA + SpikeReg + 24HMP - consistently yielded the best results for each disease type. For the healthy (GSP) data, these top two pipelines emerged as the best performers, with the composite score slightly favoring ICA-AROMA combinations, suggesting an advantage of ICA-AROMA in this context. These findings indicate that while healthy, low-motion data are relatively robust to the choice of denoising pipeline, patient datasets, especially those with higher levels of head motion, are more sensitive to the selected approach.

Overall, healthy data (GSP) consistently yielded the best results in all QC metrics, underscoring the critical role of high initial data quality in achieving effective preprocessing outcomes. While denoising techniques are designed to mitigate head motion-related artifacts and enhance signal quality, the results highlight that pipelines logically perform optimally when starting with datasets that have minimal head motion contamination. These observations suggest that, although preprocessing can significantly improve data quality, the inherent characteristics of the raw data - such as lower head motion - play a pivotal role in achieving the best outcomes for QC metrics. However, acquiring low-motion data, especially in clinical settings, is often challenging. Therefore, implementing optimized denoising strategies is essential to enhance high-motion data, elevating its quality to an acceptable and usable level where possible.

Furthermore, the choice of denoising pipeline should be closely aligned with the specific research objectives, particularly whether the study emphasizes whole-brain connectivity or fine-grained regional analyses. Consistent with our findings, pipelines incorporating ICA-AROMA (e.g., ICA + SpikeReg + 24HMP) demonstrated good performance in data from the non-lesional condition with higher head motion, suggesting them for whole-brain analyses where global noise reduction is a priority. In contrast, CC-based pipelines (e.g., CC + SpikeReg + 24HMP) excelled in data from lesional conditions, indicating their suitability for studies focusing on specific regions affected by lesions. But also, CC + SpikeReg + 24HMP managed to minimize the global noise in data form lesional conditions.

### Limitations

It is important to acknowledge several limitations. Firstly, the sample size of our study cohorts, particularly within specific subgroups such as patients in ICANS, is relatively small. This limited sample size may compromise the generalizability of our findings and could introduce biases; thus, caution is warranted when interpreting the results. Secondly, our study exclusively focuses on patients with brain tumors within the data from lesional conditions, which may constrain the generalizability regarding other forms of brain lesions, such as stroke. Another important limitation of this study is the specificity of the findings to the scanner and protocol used. The data were acquired using a specific scanner model and imaging parameters, which may limit the generalizability of these findings to other scanners and protocols. Differences in hardware, acquisition settings, and preprocessing pipelines could lead to variability in denoising efficacy and quality metrics, highlighting the need for further validation across diverse setups. Additionally, this study did not investigate potential thresholds for excluding high-motion data that is unlikely to yield meaningful FC metrics, even with the application of optimal denoising strategies. Establishing such thresholds would be critical for guiding the inclusion criteria in future studies and ensuring the reliability of results in clinical datasets. Addressing these limitations in future research endeavors would enhance our understanding of optimal denoising strategies for fcMRI data across diverse clinical populations and categories of data quality.

### Conclusion

In conclusion, our study highlights the importance of tailoring denoising strategies for fcMRI data to the specific disease type. Our findings underscore the necessity of distinguishing between data from lesional and data from non-lesional conditions when selecting denoising approaches, particularly in the context of real-world fcMRI data where head motion is prevalent. By addressing these considerations, we can ultimately enhance the reliability and utility of fcMRI as a valuable tool for investigating brain FC in patient cohorts.

### Outlook

To move beyond simply testing various denoising approaches and observing their outcomes, the next step would be to optimize the selection of fcMRI denoising strategies by employing advanced machine learning methods such as reinforcement learning, differentiable programming, and Bayesian optimization. This represents a more top-down, goal-oriented paradigm, actively learning to achieve optimal results by minimizing QC-FC correlations, QC-FC distance dependence, and tDOF-loss while maximizing connectivity within standard functional networks such as the default mode network, dynamically tailoring the denoising pipeline to the specific data and research objectives. Also, the importance of each QC-metric can then be defined based on the specific research question. Additionally, a key objective will be to establish thresholds for head motion values, which may also vary depending on the particular research questions being addressed.

## Data Availability

The data, including fcMRI scans and clinical metadata, are publicly available only for the GSP dataset, while patient cohort data remain restricted due to confidentiality and institutional privacy policies. However, de-identified patient data may be provided by the corresponding author upon reasonable request and with appropriate ethical approvals. The code used for data analysis and processing is also available upon request. For access to the code or data, please contact the corresponding author.

## Author Contributions

**Stephan Wunderlich:** Conceptualization, data curation, formal analysis, investigation, methodology, resources, supervision, validation, visualization, writing – original and final draft, review and editing. **Cagatay Alici:** Data curation, validation, visualization, writing – original draft. **Saeed Motevalli:** Data curation, validation, visualization. **Veit Stoecklein**: Data contribution, writing – review and editing final draft. **Marion Subklewe**: Data contribution, writing – review and editing final draft. **Louisa von Baumgarten**: Data contribution, writing – review and editing final draft. **Florian Schöberl**: Data contribution, writing – review and editing final draft. **Enrico Schulz:** Writing – review and editing final draft. **Sophia Stoecklein:** Supervision, MRI data curation and contribution, writing – review and editing final draft.

